# Gut-derived PPAR-γ signaling and risk of bacterial enteric infection: insight from thiazolidinedione users in a US population-based study

**DOI:** 10.1101/2024.09.24.24313682

**Authors:** Morgan Birabaharan, David C Kaelber, Victor Nizet, Amir Zarrinpar

## Abstract

**Background and Aims:** The ongoing antimicrobial resistant crisis heralds the need for new therapeutics against enteric infection. In mouse models, colon epithelial peroxisome proliferator-activated receptor-γ (PPAR-γ) signaling limits oxygen and nitrate luminal bioavailability, thereby preventing bacterial pathogen colonization. However, whether this mechanism operates similarly in humans remains uncertain.

**Methods:** To investigate, we used the cloud-based TriNetX Analytics Platform which aggregates health records from 117 million patients across 66 US healthcare organizations, to assess the risk of bacterial enteric infection among diabetic patients prescribed thiazolidinediones, a class of PPAR-γ agonists, to other anti-diabetes medications.

**Results:** Among 85,117 thiazolidinedione users, we observed a 22-49% lower risk of bacterial enteric infections compared to users of other anti-diabetes medications. This reduction in risk was consistent across high-risk individuals, regardless of sex or age. Similar results were replicated in high-risk patients when thiazolidinedione users were directly compared to those on DPP-4 inhibitors.

**Conclusion:** These findings support the potential protective role of PPAR-γ signaling against bacterial enteric infection and call for further clinical investigation.

## Introduction

The body’s primary defense against bacterial enteric infection relies heavily on the abundant and diverse intestinal microbiota [1,2]. These gut commensal microbes confer protection by direct bacteria-to-bacteria interactions and activating host immune defenses—a phenomenon known as colonization resistance [1,2]. Amid the ongoing antimicrobial resistant crisis, understanding the mechanisms of colonization resistance has gained increasing importance. Identifying microbial metabolites, proteins, or host receptors involved in preventing infection can lead to new anti-infective strategies and potentially reduce the 1.6-2.1 million annual diarrheal deaths worldwide [3].

Recent mechanistic studies have highlighted a central role of the host nuclear hormone receptor peroxisome proliferator-activated receptor (PPAR-γ) in colonization resistance [4,5]. Commensal colonic microbes, predominantly obligate anaerobes, thrive in an oxygen- and nitrate-deprived environment, limiting the expansion of facultative anaerobic organisms such as *Salmonella* and Enterobacteriaceae. These microbes maintain the low-oxygen environment by producing metabolites such as butyrate, which activates PPAR-γ in colonocytes [4,5]. This activation enhances oxygen consumption by the host, sustaining the protective anaerobic state. In mouse models, depletion of PPAR-γ signaling—triggered by antibiotics or a high-fat diet—has been associated with increased bacterial pathogenesis, aligning with well-known patient risk factors [4–9]. Pharmacological PPAR-γ agonists have also shown promising results in restoring this protective effect [10]. However, it remains unclear whether PPAR-γ offers similar protection against enteric infection in humans.

Thiazolidinediones, a class of PPAR-γ agonists, are commonly used in the management of type 2 diabetes mellitus (T2DM) [11,12]. Assessing the risk of bacterial enteric infection in thiazolidinediones users compared to those receiving other anti-diabetes medications could provide insights into the viability of targeting PPAR-γ for anti-infective purposes. In this study, we conducted a population-based analysis in the US to assess the risk of bacterial enteric infection in T2DM patients using thiazolidinediones anti-diabetes medications.

## Methods

### Data

We used the cloud-based TriNetX Analytics Platform, US Collaborative Network, to obtain web-based real-time secure access to fully deidentified electronic health records of 117 million patients from 66 health care organizations, representing 27% of the US population from all 50 states. Both inpatient and outpatient settings mostly from large academic medical institutions in the US, as well as persons from diverse geographic, age, race and ethnicity, income, and insurance groups are represented. The geographic distribution of patients from the TriNetX platform is 25% in the Northeast, 17% in the Midwest, 41% in the South, and 12% in the West, with 5% unknown.

### Ethics Statement

The TriNetX platform aggregates and HIPAA de-identifies data contributed from the electronic health records of participating healthcare organizations. The TriNetX platform also only reports population-level results (no access to individual patient data) and uses statistical blurring, reporting all population-level counts between 1 and 10 as 10. Because this study used only deidentified patient records, it was exempted from review by the MetroHealth System Institutional Review Board.

### Study populations

Risk for bacterial enteric infection can often be based on factors that are not well captured by electronic medical records, including consumption of undercooked meat or international travel, and may be a rare occurrence in those without risk factors present. Therefore, the outcome of bacterial enteric infection after prescription of thiazolidinedione or other anti-diabetes medication was determined among a “low-risk” and “high-risk” cohort. Low-risk for bacterial enteric infection was defined as no prior history of bacterial enteric infection. High-risk was defined as persons with a previous diagnosis of bacterial enteric infection, suggesting risk factors for disease such as altered gut microbiota, behavioral and environmental factors, or predisposing chronic diseases are present [13–16].

The study was designed for a 5-year recruitment date, from 3 January 2017 to an end date of 3 January 2022. The start date was chosen to help include the approval of semaglutide for T2DM management (FDA approved Dec 2017) and the end date was chosen to minimize overlap of GLP1 agonist approval for weight loss in those without diabetes (FDA approved June 2021). The index events for each were time of anti-diabetes medication prescription

### The study population at low-risk for bacterial enteric infection

Two thiazolidinediones are available in the United States, pioglitazone and rosiglitazone. Accordingly, all persons ≥18 years of age between January 2017 and January 2022 who within 1 month after a medical encounter for the diagnosis of type 2 diabetes mellitus were prescribed thiazolidinediones (pioglitazone or rosiglitazone) or non-thiazolidinediones anti-diabetes medications (Glucagon-like peptide-1 (GLP-1) analogues, insulin, metformin, sulfonylureas, alpha glucosidase inhibitors, DPP-4 inhibitors, SGLT2 inhibitors), and had no prior history of bacterial enteric infection before index event (prescription of anti-diabetes medication) were included. The study population was then divided into two cohorts: (1) thiazolidinedione-users at low risk and (2) non-thiazolidinedione users at low risk. Troglitazone users were excluded from the study as the medication is not available in the US due to its hepatoxicity. In the non-thiazolidinedione cohort, persons with thiazolidinedione prescription were excluded from the non-thiazolidinedione cohort.

### The study population at high-risk for bacterial enteric infection

Two thiazolidinediones are available in the United States, pioglitazone and rosiglitazone. Accordingly, all persons ≥18 years of age between January 2017 and January 2022 who within 1 month after a medical encounter for the diagnosis of type 2 diabetes mellitus were prescribed thiazolidinediones (pioglitazone or rosiglitazone) or non-thiazolidinediones anti-diabetes medications, and had prior history of bacterial enteric infection before index event (prescription of anti-diabetes medication) were included. The study population was then divided into two cohorts: (1) thiazolidinedione-users at high risk and (2) non-thiazolidinedione users at high risk. Troglitazone users were excluded from the study as the medication is not available in the US due to its hepatoxicity. In the non-thiazolidinedione cohort, persons with thiazolidinedione prescription were excluded from the non-thiazolidinedione cohort.

### Statistical analysis

For each study cohort, the thiazolidinedione users and non-thiazolidinedione users were propensity score matched (1:1 using nearest neighbor greedy matching with a caliper of 0.25 times the standard deviation) on covariates that are potential risk factors for bacterial enteric infection including diabetes severity, such as age, sex, race, ethnicity, overweight and obesity, lifestyle problems, ischemic heart disease, liver disease, kidney disease, lung disease, hypertension, hyperlipidemia, cerebrovascular disease, atherosclerosis, other peripheral vascular disease, tobacco use, alcohol use, human immunodeficiency virus (HIV), organ transplant, use of immunosuppressants, use of chemotherapeutics, and Hemoglobin A1C levels.

The outcome bacterial enteric infection that occurred in the 6-month time window after the index event (prescription of medication) were compared between the matched medication groups. The status of bacterial enteric infection was defined by ICD-10 A00-A05 (A00: Cholera, A01: Typhoid and paratyphoid fevers, A02: Other salmonella infections, A03: Shigellosis, A04: Other bacterial intestinal infections, A05: Other bacterial foodborne intoxications, not elsewhere classified). As counts of bacterial enteric infection were low in the cohort, risk for an individual bacterial species (i.e. *Salmonella sp*, *Enterohemorrhagic E coli*) precluded statistical analysis (most counts were less than 10, therefore were rounded to 10 by TriNetX) and was not performed. Only a composite outcome was investigated.

Several subgroup analyses were performed to assess the robustness of our results and possible bias. First, outcomes were examined when stratified by sex and age. Second, given diabetes medications are often prescribed in combination, it was necessary to evaluate whether other medications commonly administered alongside thiazolidinediones could confound observed effects. However, thiazolidinediones are rarely used for the management of T2DM because of weight gain, pedal edema, and heart failure exacerbation, leading to their classification as a third line option. Accordingly, thiazolidinediones were compared with another third-line anti-diabetes medication, dipeptidyl peptidase-4 inhibit0rs (DPP-4) inhibitors. Age and sex stratified analysis were not performed between thiazolidinediones and DPP-4 inhibitors as subgroups often had less than 10 persons with the desired outcome. TriNetX to maintain deidentification does not provide actual counts when below 10, which precludes accurate statistical analysis. All statistical analysis were performed with the TriNetX Advanced Analytics Platform. The TriNetX platform calculates RRs and associated confidence interval (CI) using R’s Survival package, version 3.2-3, with proportional hazard assumption tested using the generalized Schoenfeld approach. Statistical significance was set a 2-sided P value of <0.05.

## Results

### Association of thiazolidinedione with risk of bacterial enteric infection in patients at low risk

The study population consisted of 85,917 and 2,154,426 persons with T2DM who between 1/2017-1/2022 were prescribed thiazolidinedione or non-thiazolidinedione anti-diabetes medications, respectively, and had no prior history of bacterial enteric infection. Persons on thiazolidinediones compared to non-thiazolidinedione were more likely to be older, male, white, and had higher prevalence of overweight and obesity, chronic kidney disease, and Hgb A1C levels greater than 9 (**Table 1**).

**Table 1:**
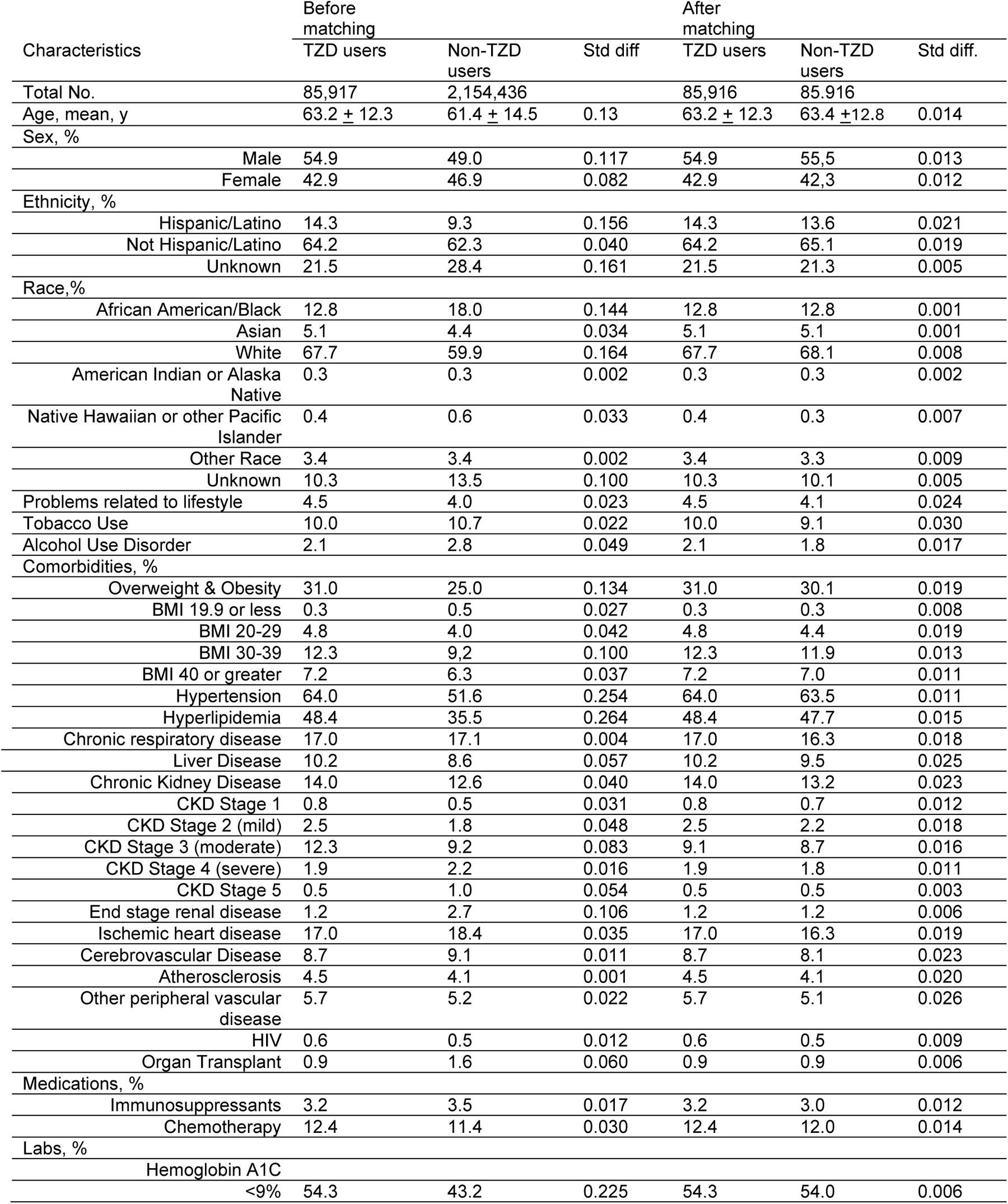
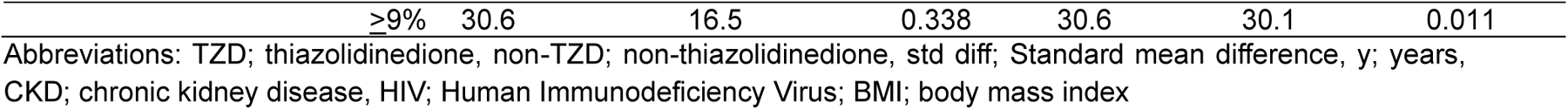
Characteristics of the thiazolidinedione cohort and the non-thiazolidinedione anti-diabetes medication cohort for the study population at low risk for bacterial enteric infection.

**Table 2:** Characteristics of the thiazolidinedione cohort and the non-thiazolidinedione anti-diabetes medication cohort for the study population at high-risk for bacterial enteric infection.

| Characteristics | Before matching |  |  | After matching |  |  |
| --- | --- | --- | --- | --- | --- | --- |
|  | TZD users | Non-TZD users | Std diff. | TZD users | Non-TZD users | Std diff. |
| Total No. | 1,975 | 57,963 |  | 1,974 | 1,974 |  |
| Age, mean, y | 63.0 ± 12.7 | 62.6 ± 14.1 | 0.029 | 63.0 ± 12.7 | 63.1 ± 13.3 | 0.007 |
| Sex, % |  |  |  |  |  |  |
| Male | 44.1 | 43.3 | 0.017 | 44.1 | 42.8 | 0.027 |
| Female | 54.3 | 52.9 | 0.029 | 54.3 | 55.1 | 0.017 |
| Ethnicity, % |  |  |  |  |  |  |
| Hispanic/Latino | 20.9 | 12.3 | 0.233 | 20.8 | 20.8 | 0.001 |
| Not Hispanic/Latino | 65.7 | 66.3 | 0.013 | 65.7 | 64.5 | 0.026 |
| Unknown | 13.5 | 21.4 | 0.211 | 13.5 | 14.7 | 0.036 |
| Race,% |  |  |  |  |  |  |
| African American/Black | 16.9 | 20.8 | 0.099 | 16.9 | 16.3 | 0.016 |
| Asian | 4.4 | 4.1 | 0.014 | 4.4 | 4.4 | <0.001 |
| White | 65.4 | 57.7 | 0.158 | 65.4 | 65.8 | 0.009 |
| American Indian or Alaska Native | 0.5 | 0.4 | 0.020 | 0.5 | 0.5 | <0.001 |
| Native Hawaiian or other Pacific Islander | 0.6 | 1.0 | 0.053 | 0.6 | 0.5 | 0.007 |
| Other Race | 3.4 | 3.3 | 0.011 | 3.4 | 3.2 | 0.011 |
| Unknown | 9.0 | 12.7 | 0.120 | 9.0 | 9.4 | 0.014 |
| Lifestyle problem | 8.6 | 7.6 | 0.036 | 8.6 | 7.4 | 0.041 |
| Tobacco Use | 16.2 | 18.2 | 0.053 | 16.2 | 14.4 | 0.049 |
| Alcohol Use Disorder | 5.3 | 6.9 | 0.067 | 5.3 | 4.4 | 0.040 |
| Comorbidities, % |  |  |  |  |  |  |
| Overweight & Obesity | 45.9 | 38.6 | 0.147 | 45.8 | 43.4 | 0.049 |
| BMI 19.9 or less | 1.8 | 2.0 | 0.011 | 1.8 | 1.6 | 0.020 |
| BMI 20-29 | 8.4 | 8.4 | 0.001 | 8.4 | 7.0 | 0.049 |
| BMI 30-39 | 17.5 | 15.2 | 0.061 | 17.5 | 16.0 | 0.039 |
| BMI 40 or greater | 12.7 | 10.5 | 0.067 | 12.6 | 11.6 | 0.031 |
| Hypertension | 79.8 | 71.5 | 0.195 | 79.8 | 77.1 | 0.067 |
| Hyperlipidemia | 67.4 | 54.7 | 0.263 | 67.4 | 64.9 | 0.051 |
| Chronic respiratory disease | 29.6 | 31.9 | 0.050 | 29.6 | 27.7 | 0.043 |
| Liver Disease | 24.4 | 21.8 | 0.061 | 24.2 | 22.2 | 0.052 |
| Chronic Kidney Disease | 28.7 | 32.5 | 0.084 | 28.6 | 27.4 | 0.027 |
| CKD Stage 1 | 2.0 | 1.5 | 0.036 | 1.9 | 2.3 | 0.028 |
| CKD Stage 2 (mild) | 5.7 | 5.1 | 0.026 | 5.7 | 6.3 | 0.026 |
| CKD Stage 3 (moderate) | 19.3 | 18.2 | 0.029 | 19.3 | 18.2 | 0.027 |
| CKD Stage 4 (severe) | 5.4 | 8.6 | 0.125 | 5.4 | 5.1 | 0.014 |
| CKD Stage 5 | 1.7 | 5.0 | 0.184 | 1.7 | 1.6 | 0.012 |
| End stage renal disease | 4.4 | 10.9 | 0.248 | 4.4 | 4.0 | 0.020 |
| Ischemic heart disease | 28.7 | 33.9 | 0.113 | 28.7 | 26.2 | 0.055 |
| Cerebrovascular Disease | 17.0 | 18.9 | 0.050 | 16.9 | 14.6 | 0.064 |
| Atherosclerosis | 10.9 | 12.2 | 0.043 | 10.9 | 9.2 | 0.056 |
| Other peripheral vascular disease | 11.6 | 12.8 | 0.037 | 11.6 | 10.0 | 0.051 |
| HIV | 1.4 | 1.3 | 0.007 | 1.4 | 1.3 | 0.009 |
| Organ Transplant | 3.7 | 8.0 | 0.184 | 3.7 | 4.0 | 0.013 |
| Medications |  |  |  |  |  |  |
| Immunosuppressants | 8.2 | 11.7 | 0.116 | 8.2 | 7.0 | 0.044 |
| Chemotherapy | 24.8 | 25.6 | 0.020 | 24.8 | 22.7 | 0.049 |
| Labs |  |  |  |  |  |  |
| Hemoglobin A1C |  |  |  |  |  |  |
| <9% | 69.8 | 61.6 | 0.173 | 69.8 | 69.1 | 0.014 |
| ≥9% | 43.0 | 24.5 | 0.400 | 43.0 | 43.2 | 0.003 |

Abbreviations: TZD; thiazolidinedione, non-TZD; non-thiazolidinedione, std diff; Standard mean difference, y; years, CKD; chronic kidney disease, HIV; Human Immunodeficiency Virus; BMI; body mass index

After 1:1 propensity matching, demographic and clinical characteristics were balanced (85,916 in each cohort, 63.2 years of age, 42.9% Female, 12.8% Black, 67.7% White, 14.3% Hispanic) (**Table 1**). Matched cohorts were followed for 6 months after the index event. Compared to anti-diabetes non-thiazolidinedione medications, thiazolidinedione was associated with a lower risk of incident bacterial enteric infection among those at low-risk (0.03% vs 0.06%; RR 0.51, 95% CI 0.32, 0.82). An association of lower risk for bacterial enteric infection was observed across age and sex, however only among those >65 years were findings statistically significant (0.02% vs 0.05%; RR 0.44, 95% CI 0.23,0.88) (**Figure 1**).

**Figure 1:**
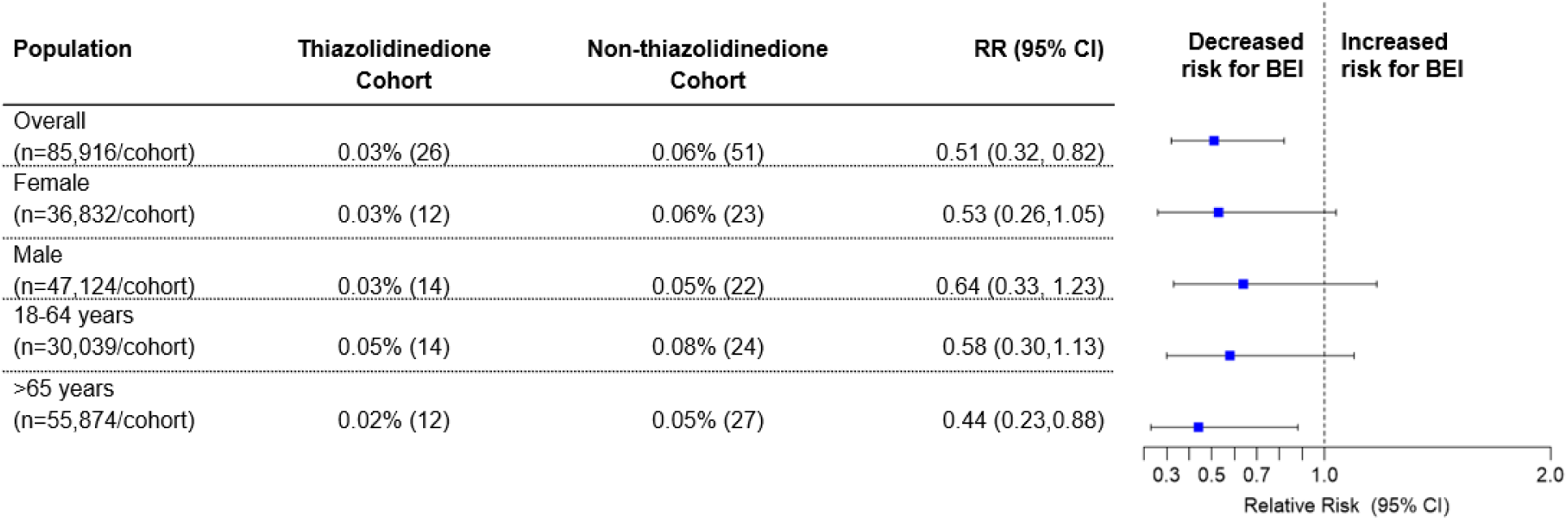
Risk of bacterial enteric infection in low-risk individuals (no prior history of bacterial enteric infection); BEI, Bacterial Enteric Infection

### Association of thiazolidinedione with risk of bacterial enteric infection in patients at high risk

The study population consisted of 1,975 and 57,963 persons with T2DM who between 1/2017-1/2022 were prescribed thiazolidinedione or non-thiazolidinedione anti-diabetes medications, respectively, and had prior history of bacterial enteric infection. Persons on thiazolidinediones were more likely to be Hispanic and White, and had higher prevalence of overweight and obesity, hypertension, hyperlipidemia, and Hb1C levels greater than 9.

After 1:1 propensity matching, demographic and clinical characteristics were balanced (1,974 in each cohort, 63.0 years of age, 44.1% Female, 16.9% Black, 65.4% White, 20.8% Hispanic) (**Table 1**). Matched cohorts were followed for 6 months after the index event. Compared to anti-diabetes non-thiazolidinedione medications, thiazolidinedione was associated with a lower risk of incident bacterial enteric infection among those at high risk (17.6% vs 22.5%; RR 0.78, 95% CI 0.69,0.88). An association of lower risk for bacterial enteric infection was observed across all sex and age subgroups (**Figure 2**).

**Figure 2:**
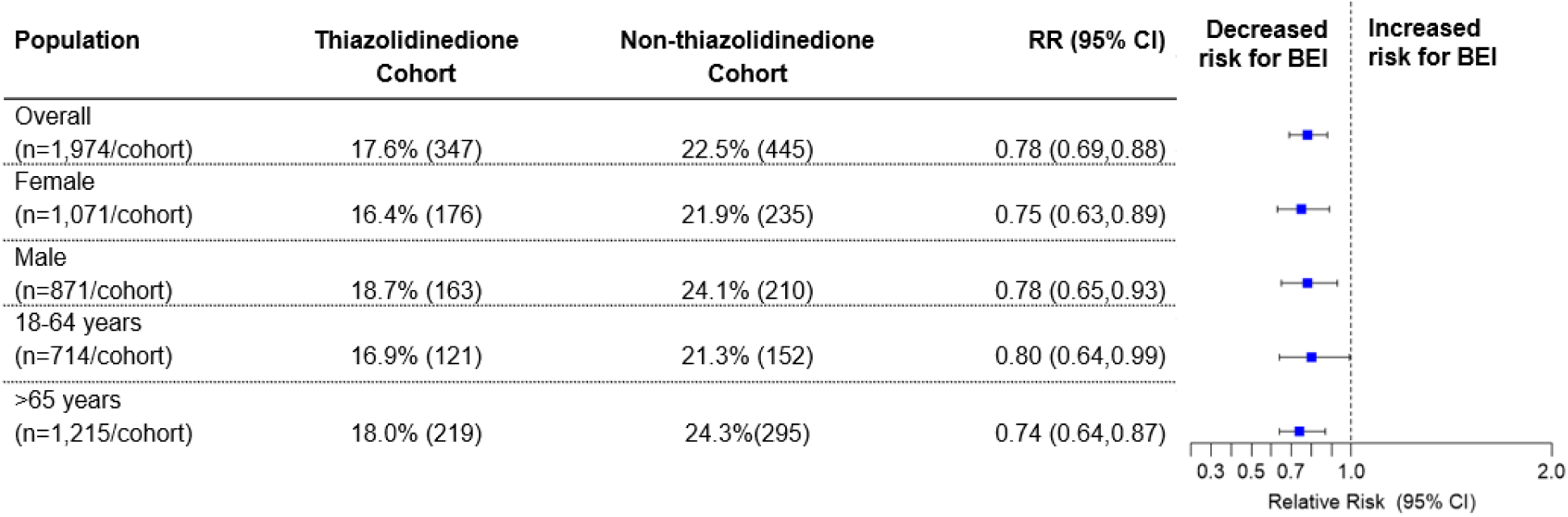
Risk of bacterial enteric infection in high-risk individuals (prior history of bacterial enteric infection); BEI, Bacterial Enteric Infection

### Association of thiazolidinedione compared to DPP-4 inhibitors with risk of bacterial enteric infection in patients at low and high risk (sensitivity analysis)

In the matched cohorts, thiazolidinedione use compared to those on DPP-4 inhibitors in patients with T2DM was associated with a lower risk of bacterial enteric infection in those at high-risk (RR 0.82, 95% CI 0.72,0.93) and not those at low-risk (RR 0.96, 95% CI 0.56, 1.65) (**Figure 3**).

**Figure 3:**
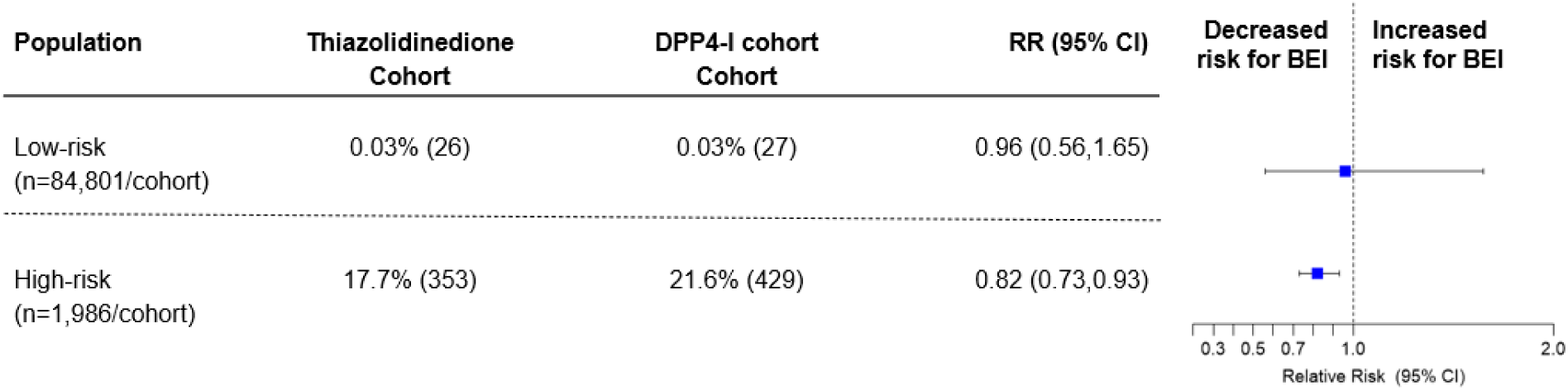
Risk of bacterial enteric infection in thiazolidine users compared to DPP-4 inhibitors with T2DM in low-risk (no prior history of bacterial enteric infection) and high-risk (prior history of bacterial enteric infection); BEI, Bacterial Enteric Infection, T2DM, Type 2 Diabetes Mellitus

## Discussion

Our analysis identified a significant association between thiazolidinedione use and a reduced risk of bacterial enteric infections in T2DM patients, consistent with findings from animal models. This protective effect was observed in both low- and high-risk populations, with notable reductions in infection across various age and sex subgroups, particularly in high-risk individuals with a history of prior infections. The reduced risk seen when comparing thiazolidinediones to third-line diabetes medications (DPP-4 inhibitors) further supports the hypothesis that modulating PPAR-γ signaling mitigate bacterial enteric infections, especially in high-risk situations—for example, protracted antibiotic use during hospitalization, international travel to high incidence locales, or outbreaks in nursing homes and chronic care facilities.

Thiazolidinediones are typically used as a third-line medications when other anti-diabetes medications have proven ineffective or when insurance limitations restrict alternative options [12]. Consequently, thiazolidinedione users often represent a population with more severe or poorly controlled diabetes, characterized by frequent healthcare encounters and lower socioeconomic status [17–18]. These factors are associated with an increased risk of bacterial enteric infection, suggesting the protective effect of thiazolidinediones may be greater than what this analysis indicates [19–21].

Repurposing thiazolidinediones is not a novel concept. Initially, PPAR-γ was believed to primarily function on adipose tissue, regulating insulin resistance, which led to the development of thiazolidinediones for diabetes management [22]. However, investigators soon appreciated the abundance of PPAR-γ in colonic epithelial cells and its involvement in colon cancer [23,24]. Subsequently, PPAR-γ signaling was shown to reduce colonic inflammation, prompting the exploration of thiazolidinediones for inflammatory bowel disease [25]. Promising results were observed in mouse models and clinical trials [25–27]. Despite these benefits, concerns over toxicities, such as weight gain, fluid retention, and heart failure, have overshadowed the benefits of thiazolidinediones. Therefore, while this study suggests thiazolidinediones may protect against bacterial enteric infection, their repurposing for this indication is likely limited by their associated risks.

Fortunately, there are alternative ways to increase PPAR-γ signaling. For instance, mesalamine (5-ASA), a PPAR-γ agonist widely used for treating mild to moderate ulcerative colitis, may offer a safer alternative. Studies suggest that 5-ASA prevents the expansion of colitogenic bacteria such as *Escherichia coli* [10], indicating its potential role in preventing bacterial enteric infection in high-risk individuals.

Dietary modifications may also enhance PPAR-y signaling. A high-fiber diet can increase butyrate levels, which, in turn, activate PPAR-y. However, this depends on the presence of a gut microbiota capable of butyrate production. Unsaturated fatty acids, found in dairy and meat products, are natural PPAR-y ligands and may also promote colonic health, although achieving sufficient intraluminal concentrations *in vivo* remains controversial. Small animal studies have demonstrated possible benefits from such dietary interventions [28,29].

One hypothesis behind the protective effects of PPAR-γ activation is its ability to maintain an anaerobic environment in the gut, which helps prevent bacterial enteric infections. However, this mechanism does not directly explain why thiazolidinediones may also protect against *Clostridioides difficile*, an obligate anaerobe. It is possible that PPAR-γ signaling supports the growth of beneficial anaerobes such as *Clostridium scindens* or *Blautia producta*, which inhibit *C. difficile* through the production of secondary bile acids and lantibiotics, respectively [30,31]. Further, PPAR-γ activity in macrophages, which has been shown to ameliorate colonic inflammation, may also contribute to this protective effect. Given that thiazolidinediones are primarily absorbed in the small intestine and may not directly reach the colon, this macrophage-mediated mechanism merits further investigation [10].

## Limitations

There are limitations that bear consideration when interpreting the results of this study. Residual confounding, particularly diabetes severity, may impact the findings. Although we controlled for factors like ischemic heart disease, peripheral vascular disease, chronic kidney disease, and HbA1C levels, fully characterizing the severity of diabetes remains a challenge. Further, medication adherence cannot be confirmed based on electronic health records. However, our inclusion of individuals with thiazolidinedione or other anti-diabetes prescriptions following a healthcare encounter for T2DM suggests active management of the disease. Limited sample sizes precluded an analysis of dose-dependent effects and we could not assess the duration of thiazolidinedione use required for a protective effect. Further, since T2DM is often managed with a combination of medications, it is possible that other medications commonly used alongside thiazolidinediones contributed to the observed protective effect. To address this, we conducted a direct comparison with DPP-4 inhibitors, which showed a decreased risk of bacterial enteric infection in high-risk individuals. Lastly, the severity of enteric infections was not captured in this study, so it remains unknown whether thiazolidinediones can reduce disease severity.

## Conclusion

Bacterial enteric infections remain a significant public health concern, with acute gastroenteritis causing an estimated 179 million outpatient visits and nearly 500,000 hospitalizations annually [32–33]. Current antimicrobial treatments are often inadequate, as antibiotics can prolong symptoms, increase bacterial shedding, and lead to recurrence [14–16]. Given the limitations of existing treatments, preserving beneficial intestinal microbes with alternative therapies that minimize collateral damage to the microbiome is a key consideration. In this context PPAR-γ agonists may offer a promising alternative.

## Data Availability

This study used population-level aggregate and deidentified data generated by the TriNetX Platform. Due to data privacy, patient-level data was not used and cannot be shared

## ACKNOWLEDGMENTS

MB is supported by T32 AI007036. DCK is supported by the Clinical and Translational Science Collaborative of Northern Ohio which is funded by the National Center for Advancing Translational Sciences (NCATS) of the National Institutes of Health, UM1TR004528. AZ is supported by NIH R01AZ is supported by NIH R01 EB030134, R01 AI163483, U01 CA265719, and VA Merit BLR&D Award I01 BX005707. All authors receive institutional support from NIH P30 DK120515, P30 DK063491, and UL1 TR001442.

## References

1. Caballero-Flores G, Pickard JM, Núñez G. Microbiota-mediated colonization resistance: mechanisms and regulation. Nat Rev Microbiol. 2023 Jun;21(6):347–360. doi: 10.1038/s41579-022-00833-7. Epub 2022 Dec 20. PMID: 36539611; PMCID: PMC10249723.

2. Buffie CG, Pamer EG. Microbiota-mediated colonization resistance against intestinal pathogens. Nat Rev Immunol. 2013 Nov;13(11):790–801. doi: 10.1038/nri3535. Epub 2013 Oct 7. PMID: 24096337; PMCID: PMC4194195.

3. Petri WA Jr, Miller M, Binder HJ, Levine MM, Dillingham R, Guerrant RL. Enteric infections, diarrhea, and their impact on function and development. J Clin Invest. 2008 Apr;118(4):1277–90. doi: 10.1172/JCI34005. PMID: 18382740; PMCID: PMC2276781.

4. Byndloss MX, Olsan EE, Rivera-Chávez F, Tiffany CR, Cevallos SA, Lokken KL, Torres TP, Byndloss AJ, Faber F, Gao Y, Litvak Y, Lopez CA, Xu G, Napoli E, Giulivi C, Tsolis RM, Revzin A, Lebrilla CB, Bäumler AJ. Microbiota-activated PPAR-γ signaling inhibits dysbiotic Enterobacteriaceae expansion. Science. 2017 Aug 11;357(6351):570-575. doi: 10.1126/science.aam9949. PMID: 28798125; PMCID: PMC5642957.

5. Litvak Y, Byndloss MX, Bäumler AJ. Colonocyte metabolism shapes the gut microbiota. Science. 2018 Nov 30;362(6418):eaat9076. doi: 10.1126/science.aat9076. PMID: 30498100; PMCID: PMC6296223.

6. Yoo W, Zieba JK, Foegeding NJ, Torres TP, Shelton CD, Shealy NG, Byndloss AJ, Cevallos SA, Gertz E, Tiffany CR, Thomas JD, Litvak Y, Nguyen H, Olsan EE, Bennett BJ, Rathmell JC, Major AS, Bäumler AJ, Byndloss MX. High-fat diet-induced colonocyte dysfunction escalates microbiota-derived trimethylamine *N*-oxide. Science. 2021 Aug 13;373(6556):813-818. doi: 10.1126/science.aba3683. PMID: 34385401; PMCID: PMC8506909.

7. Lee JY, Cevallos SA, Byndloss MX, Tiffany CR, Olsan EE, Butler BP, Young BM, Rogers AWL, Nguyen H, Kim K, Choi SW, Bae E, Lee JH, Min UG, Lee DC, Bäumler AJ. High-Fat Diet and Antibiotics Cooperatively Impair Mitochondrial Bioenergetics to Trigger Dysbiosis that Exacerbates Pre-inflammatory Bowel Disease. Cell Host Microbe. 2020 Aug 12;28(2):273–284.e6. doi: 10.1016/j.chom.2020.06.001. Epub 2020 Jul 14. PMID: 32668218; PMCID: PMC7429289.

8. Rivera-Chávez F, Zhang LF, Faber F, Lopez CA, Byndloss MX, Olsan EE, Xu G, Velazquez EM, Lebrilla CB, Winter SE, Bäumler AJ. Depletion of Butyrate-Producing Clostridia from the Gut Microbiota Drives an Aerobic Luminal Expansion of Salmonella. Cell Host Microbe. 2016 Apr 13;19(4):443–54. doi: 10.1016/j.chom.2016.03.004. PMID: 27078066; PMCID: PMC4832419.

9. Shelton CD, Sing E, Mo J, Shealy NG, Yoo W, Thomas J, Fitz GN, Castro PR, Hickman TT, Torres TP, Foegeding NJ, Zieba JK, Calcutt MW, Codreanu SG, Sherrod SD, McLean JA, Peck SH, Yang F, Markham NO, Liu M, Byndloss MX. An early-life microbiota metabolite protects against obesity by regulating intestinal lipid metabolism. Cell Host Microbe. 2023 Oct 11;31(10):1604–1619.e10. doi: 10.1016/j.chom.2023.09.002. Epub 2023 Oct 3. PMID: 37794592; PMCID: PMC10593428.

10. Cevallos SA, Lee JY, Velazquez EM, Foegeding NJ, Shelton CD, Tiffany CR, Parry BH, Stull-Lane AR, Olsan EE, Savage HP, Nguyen H, Ghanaat SS, Byndloss AJ, Agu IO, Tsolis RM, Byndloss MX, Bäumler AJ. 5-Aminosalicylic Acid Ameliorates Colitis and Checks Dysbiotic Escherichia coli Expansion by Activating PPAR-γ Signaling in the Intestinal Epithelium. mBio. 2021 Jan 19;12(1):e03227–20. doi: 10.1128/mBio.03227-20. PMID: 33468700; PMCID: PMC7845635.

11. Yki-Järvinen H. Thiazolidinediones. N Engl J Med. 2004 Sep 9;351(11):1106–18. doi: 10.1056/NEJMra041001. PMID: 15356308.

12. 12. American Diabetes Association Professional Practice Committee. 9. Pharmacologic Approaches to Glycemic Treatment: Standards of Medical Care in Diabetes-2022. Diabetes Care. 2022 Jan 1;45(Suppl 1):S125–S143. doi: 10.2337/dc22-S009. PMID: 34964831.

13. Shane AL, Mody RK, Crump JA, Tarr PI, Steiner TS, Kotloff K, Langley JM, Wanke C, Warren CA, Cheng AC, Cantey J, Pickering LK. 2017 Infectious Diseases Society of America Clinical Practice Guidelines for the Diagnosis and Management of Infectious Diarrhea. Clin Infect Dis. 2017 Nov 29;65(12):e45–e80. doi: 10.1093/cid/cix669. PMID: 29053792; PMCID: PMC5850553.

14. Humphries RM, Linscott AJ. Practical Guidance for Clinical Microbiology Laboratories: Diagnosis of Bacterial Gastroenteritis. Clin Microbiol Rev. 2015 Jan;28(1):3–31. doi: 10.1128/CMR.00073-14. PMID: 25567220; PMCID: PMC4284301.

15. Peniche AG, Savidge TC, Dann SM. Recent insights into Clostridium difficile pathogenesis. Curr Opin Infect Dis. 2013 Oct;26(5):447–53. doi: 10.1097/01.qco.0000433318.82618.c6. PMID: 23982235.

16. Crump JA, Sjölund-Karlsson M, Gordon MA, Parry CM. Epidemiology, Clinical Presentation, Laboratory Diagnosis, Antimicrobial Resistance, and Antimicrobial Management of Invasive Salmonella Infections. Clin Microbiol Rev. 2015 Oct;28(4):901–37. doi: 10.1128/CMR.00002-15. PMID: 26180063; PMCID: PMC4503790.

17. Arnold SV, Inzucchi SE, Echouffo-Tcheugui JB, Tang F, Lam CSP, Sperling LS, Kosiborod M. Understanding Contemporary Use of Thiazolidinediones. Circ Heart Fail. 2019 Jun;12(6):e005855. doi: 10.1161/CIRCHEARTFAILURE.118.005855. PMID: 31129998.

18. Qato DM, Trivedi AN, Mor V, Dore DD. Disparities in Discontinuing Rosiglitazone Following the 2007 FDA Safety Alert. Med Care. 2016 Apr;54(4):406–13. doi: 10.1097/MLR.0000000000000502. PMID: 26978569; PMCID: PMC4807625.

19. Chang M, Groseclose SL, Zaidi AA, Braden CR. An ecological analysis of sociodemographic factors associated with the incidence of salmonellosis, shigellosis, and E. coli O157:H7 infections in US counties. Epidemiol Infect. 2009 Jun;137(6):810–20. doi: 10.1017/S0950268808001477. Epub 2008 Oct 24. PMID: 18947443.

20. Thaiss CA, Levy M, Grosheva I, Zheng D, Soffer E, Blacher E, Braverman S, Tengeler AC, Barak O, Elazar M, Ben-Zeev R, Lehavi-Regev D, Katz MN, Pevsner-Fischer M, Gertler A, Halpern Z, Harmelin A, Aamar S, Serradas P, Grosfeld A, Shapiro H, Geiger B, Elinav E. Hyperglycemia drives intestinal barrier dysfunction and risk for enteric infection. Science. 2018 Mar 23;359(6382):1376-1383. doi: 10.1126/science.aar3318. Epub 2018 Mar 8. PMID: 29519916.

21. Bobo LD, Dubberke ER. Recognition and prevention of hospital-associated enteric infections in the intensive care unit. Crit Care Med. 2010 Aug;38(8 Suppl):S324-34. doi: 10.1097/CCM.0b013e3181e69f05. PMID: 20647790; PMCID: PMC3657464.

22. Dubuquoy L, Rousseaux C, Thuru X, Peyrin-Biroulet L, Romano O, Chavatte P, Chamaillard M, Desreumaux P. PPARgamma as a new therapeutic target in inflammatory bowel diseases. Gut. 2006 Sep;55(9):1341–9. doi: 10.1136/gut.2006.093484. PMID: 16905700; PMCID: PMC1860011.

23. Saez E, Tontonoz P, Nelson MC, Alvarez JG, Ming UT, Baird SM, Thomazy VA, Evans RM. Activators of the nuclear receptor PPARgamma enhance colon polyp formation. Nat Med. 1998 Sep;4(9):1058–61. doi: 10.1038/2042. PMID: 9734400.

24. Sarraf P, Mueller E, Jones D, King FJ, DeAngelo DJ, Partridge JB, Holden SA, Chen LB, Singer S, Fletcher C, Spiegelman BM. Differentiation and reversal of malignant changes in colon cancer through PPARgamma. Nat Med. 1998 Sep;4(9):1046–52. doi: 10.1038/2030. PMID: 9734398.

25. Su CG, Wen X, Bailey ST, Jiang W, Rangwala SM, Keilbaugh SA, Flanigan A, Murthy S, Lazar MA, Wu GD. A novel therapy for colitis utilizing PPAR-gamma ligands to inhibit the epithelial inflammatory response. J Clin Invest. 1999 Aug;104(4):383–9. doi: 10.1172/JCI7145. PMID: 10449430; PMCID: PMC408529.

26. Lewis JD, Lichtenstein GR, Stein RB, Deren JJ, Judge TA, Fogt F, Furth EE, Demissie EJ, Hurd LB, Su CG, Keilbaugh SA, Lazar MA, Wu GD. An open-label trial of the PPAR-gamma ligand rosiglitazone for active ulcerative colitis. Am J Gastroenterol. 2001 Dec;96(12):3323–8. doi: 10.1111/j.1572-0241.2001.05333.x. PMID: 11774944.

27. Lewis JD, Lichtenstein GR, Deren JJ, Sands BE, Hanauer SB, Katz JA, Lashner B, Present DH, Chuai S, Ellenberg JH, Nessel L, Wu GD; Rosiglitazone for Ulcerative Colitis Study Group. Rosiglitazone for active ulcerative colitis: a randomized placebo-controlled trial. Gastroenterology. 2008 Mar;134(3):688–95. doi: 10.1053/j.gastro.2007.12.012. Epub 2007 Dec 7. PMID: 18325386; PMCID: PMC2276587.

28. Hontecillas R, Wannemeulher MJ, Zimmerman DR, Hutto DL, Wilson JH, Ahn DU, Bassaganya-Riera J. Nutritional regulation of porcine bacterial-induced colitis by conjugated linoleic acid. J Nutr. 2002 Jul;132(7):2019–27. doi: 10.1093/jn/132.7.2019. PMID: 12097686.

29. Bassaganya-Riera J, Reynolds K, Martino-Catt S, Cui Y, Hennighausen L, Gonzalez F, Rohrer J, Benninghoff AU, Hontecillas R. Activation of PPAR gamma and delta by conjugated linoleic acid mediates protection from experimental inflammatory bowel disease. Gastroenterology. 2004 Sep;127(3):777–91. doi: 10.1053/j.gastro.2004.06.049. PMID: 15362034.

30. Buffie CG, Bucci V, Stein RR, McKenney PT, Ling L, Gobourne A, No D, Liu H, Kinnebrew M, Viale A, Littmann E, van den Brink MR, Jenq RR, Taur Y, Sander C, Cross JR, Toussaint NC, Xavier JB, Pamer EG. Precision microbiome reconstitution restores bile acid mediated resistance to Clostridium difficile. Nature. 2015 Jan 8;517(7533):205-8. doi: 10.1038/nature13828. Epub 2014 Oct 22. PMID: 25337874; PMCID: PMC4354891.

31. Kim SG, Becattini S, Moody TU, Shliaha PV, Littmann ER, Seok R, Gjonbalaj M, Eaton V, Fontana E, Amoretti L, Wright R, Caballero S, Wang ZX, Jung HJ, Morjaria SM, Leiner IM, Qin W, Ramos RJJF, Cross JR, Narushima S, Honda K, Peled JU, Hendrickson RC, Taur Y, van den Brink MRM, Pamer EG. Microbiota-derived lantibiotic restores resistance against vancomycin-resistant Enterococcus. Nature. 2019 Aug;572(7771):665-669. doi: 10.1038/s41586-019-1501-z. Epub 2019 Aug 21. PMID: 31435014; PMCID: PMC6717508.

32. Scallan E, Griffin PM, Angulo FJ, Tauxe RV, Hoekstra RM. Foodborne illness acquired in the United States--unspecified agents. Emerg Infect Dis. 2011 Jan;17(1):16–22. doi: 10.3201/eid1701.091101p2. PMID: 21192849; PMCID: PMC3204615.

33. Kendall ME, Crim S, Fullerton K, Han PV, Cronquist AB, Shiferaw B, Ingram LA, Rounds J, Mintz ED, Mahon BE. Travel-associated enteric infections diagnosed after return to the United States, Foodborne Diseases Active Surveillance Network (FoodNet), 2004-2009. Clin Infect Dis. 2012 Jun;54 Suppl 5:S480–7. doi: 10.1093/cid/cis052. PMID: 22572673.

